# Measuring the strength of maternal, newborn and child health care implementation and its association with childhood mortality risk in three rural districts of Tanzania

**DOI:** 10.1101/2023.05.10.23289812

**Authors:** Colin Baynes, Almamy Malick Kanté, Amon Exavery, Tani Kassimu, Gloria Sikustahili, Hildegalda Mushi, Kate Ramsey, Kenneth Sherr, Bryan Weiner, James F. Phillips

**Author notes:** CB conceptualized the study, had management and coordination responsibility for the data collection, developed the analytic methodology, led the analysis of data, and wrote the original and final draft of the paper. AMK and KR had oversight and leadership responsibility for the research activity, reviewed and edited the paper. AE and TK curated the data used for this analysis, participated in data collection, and reviewed and edited the paper. GS and HM participated in data collection, and reviewed and edited the paper. KS and BW validated the reproducibility of the results of the analysis, reviewed and edited the paper. JFP acquired financial support for this study, validated the reproducibility of the results of the analysis, reviewed and edited the paper.

## Abstract

This observational cohort study explores the association between maternal, newborn and child health care implementation strength and child survival in rural Tanzania from 2011-2015. We used data from a 2011 service availability and readiness assessment that quantified primary health care facilities’ ability to implement maternal, newborn and child health services and a population-level household survey that measured the utilization of such services to develop domain-specific summary measures of the effective coverage of facility-based maternal, newborn and child health care. We reduced domain specific effective coverage scores into fewer, independent scales of implementation strength using principal components analysis, and integrated them into gradients of the collective implementation strength exerted by groups of facilities on villages they served using Bayesian mixed effects models. We linked these scales to longitudinal data on the survival of children that were born in the catchment areas of the surveyed health facilities during the assessment period and followed up until December 31, 2015. We fit survival time models to estimate the relationship between implementation strength and child mortality. Increases in the implementation strength gauged by our first scale, which represented general facility readiness and the provision of antenatal, postnatal, and early childhood preventive services, were associated with child mortality risks that were, on average, 0.62 times lower. Increases in implementation strength gauged by our second scale, which represented sick childcare service provision, were associated with child mortality risks that were, on average, 0.56 times lower. We detected no significant child mortality response to our third scale, which represented intrapartum care provision. The findings suggest that strong implementation of antenatal, postnatal, early childhood preventive services and sick child care can accelerate child mortality reduction and that routine data on service availability and readiness can be used to measure health systems strengthening and its impacts.

## Introduction

In the past two decades, the spread of evidence-based child survival interventions has precipitated large-scale reductions in child mortality globally; however, considerable geographic disparities exist (1,2). Preventable loss of life during childhood remains concentrated in sub-Saharan Africa (sSA), where children are between six and fifteen times more likely to die before reaching age five compared to children in more developed regions (3,4). To address this inequality, health sectors in sSA have departed disease-specific programming strategies and adopted holistic frameworks for bundling low cost, evidence-based interventions (EBI) along the maternal, newborn and child health (MNCH) ‘continuum of care’, and integrated these bundles into primary health care (PHC) delivery systems (5–8). This introduced challenges to the task of assessing the impact of child survival programs. Whereas evaluations had traditionally focused on the effectiveness of specific interventions, the advent of new programming approaches implied the need to measure the ‘implementation strength’ with which PHC systems deliver EBI packages and evaluate whether improvement in implementation strength is associated with child survival (9–12).

The term ‘implementation strength’ has been used interchangeably with other terms, such as ‘implementation intensity’ and ‘effective coverage’ and is defined here as a quantitative measure of the ‘dose’, or the amount of input or activity, delivered to implement a program. Implementation strength has a rich history in global health (13,14). Efforts to scale up emergency obstetric care led to a ‘signal function framework’, which acknowledged that a set of performance indicators was needed to evaluate maternal survival programs since the occurrence of obstetric emergencies was rare within program settings (15). For similar reasons, researchers use signal functions to gauge the effects of expanding access to abortion care services (16,17). The advent of ‘health systems strengthening’ focused attention on the need to measure processes and outcomes of improving the capacity of health systems to deliver PHC (18,19).The World Health Organization’s ‘health systems building blocks’ framework was central to this and provided a structure for monitoring and evaluating health systems performance (20–22). This has included large-scale health facility and household surveys in low- and middle-income countries, notably Service Provision Assessments, Demographic and Health Surveys (DHS), Service Availability and Readiness Assessments (SARA), and Multiple Indicator Cluster Surveys (23–25). In addition, researchers have adapted common evaluation frameworks and applied them across health systems to measure implementation strength (26,27). Approaches, such as the Balanced Score Card, provide a dashboard of performance indicators that are used to detect variation and change in PHC implementation strength (28–32). Similar approaches have been adapted to measure the same with respect to domains of care, such as emergency obstetric and newborn care (EmONC) (33,34), integrated management of childhood illness (IMCI) (12), and family planning (35).

### Implementation strength: Measurement and association with child health gains

Efforts to apply the concept of implementation strength and evaluate its child health impacts have faced methodological challenges. The first relates to derivation of the exposure variable. A common approach has been to presuppose the intervention components that are most crucial to the causal pathway and their relative importance based on intimate knowledge of the program, literature review and credence to global measurement approaches (36,37). Researchers then design methods and tools for capturing data on those components and perform simple or weighted additive analysis to generate summary indices that represent implementation strength. Studies that have used this approach to measure the individual-level child health response to implementation strength have reported that there is a positive association between ‘dose strength’ of EBI implementation and desirous MNCH behaviors (38,39). Although their exposure measurement strategy is easy to conduct, theoretically justifiable and can produce results amenable to comparison between geographies, it assumes that implementation strength is unidimensional, and can create non-normal, skewed distributions. Thus, the additive summary score is not always conceptually meaningful and may not accurately portray overall implementation strength (40,41). Furthermore, this approach does not consider patterns between the wider, underlying set of variables of program inputs and activities between units of analysis (e.g., facility, district). This is problematic since these relationships contribute to the real variation between analytic units in terms of the underlying construct that represents the exposure of interest, i.e., the implementation strength that the health system exerted during the time and place of the evaluation at hand (42). Neglecting this issue risks misallocating weight to given input and activity variables and omitting potentially crucial ones from analysis (43). This, in turn, may undermine the quality of associational analyses which assume that differences in measures of implementation explain variation in health outcome.

Second, several analyses of the relationship between implementation strength and child health have aggregated data on outcomes to the level of district, country or other clusters that represent the level at which implementation actions are taken. Such studies have generated conflicting results with respect to the association between dose strength of program implementation and child health related care-seeking and child mortality (39,44). While this may be due to differences in the strategy and content of programs, it’s important to recognize that these studies are beset with ecological biases (45). For example, one study in Uganda, which compared approaches for linking individual-level service utilization indicators from household surveys with data on the provision of maternal and newborn services at the facility- and ecological-level (district) to estimate levels of effective coverage, found large discrepancies between estimates between the two approaches, concluding that the ecological approach appreciably overestimates effective coverage (46). Third, individual and ecological studies of implementation strength and child health have almost exclusively depended on cross-sectional data (38,39,44). Although this often reflects the best use of data available to address the research question, it precludes ascertainment of temporal order between exposure to program intensity and the occurrence of the outcome. This temporal bias raises questions about ‘reverse causality’ and prevents a ‘dose response’ interpretation of the relationship between implementation strength and child health gains.

### Purpose and objectives

The purpose of this paper is to demonstrate an analytic approach that overcomes these challenges. We first develop summary indices of implementation strength using data from a SARA that was conducted in three rural districts of Tanzania between May and August of 2011. SARA are comprised of measures of general and domain specific attributes of PHC facilities’ structural quality and are frequently implemented in low- and middle-income countries by special projects or to assess health sector performance at national scale (47). Our analysis blends weighted-additive methods and multivariate statistical procedures and applies them on a voluminous range of indicators on MNCH provision. In doing so, we derive a minimal set of independent scales that collectively represent variation in the intensity with which local health systems in the three districts provided MNCH EBI. The analysis proceeds by linking scales of implementation strength with longitudinal data that contain the survival trajectories of approximately 9,000 children, starting at their birth from March to November 2011 in the catchment villages served by SARA facilities, until the end of the cohort in December 2015. With this, we assess whether differences in the dose of implementation strength exerted by PHC facilities on villages in which children experienced infancy are associated with these children’s risk of dying during childhood.

In conducting this analysis, we address two objectives. The first is to validate the methodology we use to measure implementation strength using routine facility data obtained through a popular data collection methodology. We hypothesize that detection of an association between implementation strength and child mortality, given that the SARA measured the provision of child survival EBI that have been proven repeatedly and in various contexts, indicates that our measurement approach is worthy of consideration for uptake by other studies that use SARA or similar data to evaluate health system performance. Our second objective is to identify the components of the Tanzanian integrated PHC program that drove child mortality reduction and give insight on inputs and activities to emphasize to maximize program impact on child survival.

## Materials and Methods

### The Tanzanian health system

Since its founding, Tanzania has demonstrated commitment to achieving universal access to PHC. In the 1970s, the country launched a national expansion of PHC facilities, called dispensaries and rolled out a national village health worker program (48). During this period, the Ministry of Health promulgated a policy that guaranteed mothers and children free access to basic services such as immunization, nutrition, antenatal and postnatal care, growth monitoring, and treatment for minor illness (49). In 2007, the Ministry of Health initiated the Primary Health Care Services Improvement Program, which accelerated the expansion of dispensaries and higher-level PHC facilities, called health centers, across the country (50). Since then, strategies to guide implementation of this policy have emphasized making the above essential service package, as well as skilled obstetric care, the IMCI, and family planning available at the health center and dispensary level (51–55). The implementation of these strategies co-occurred with reduction in child mortality in Tanzania, which between 2000 and 2015 declined from 130 to 58 per 1,000 live births (56).

### Study environment and data sources

Data for this study were obtained from the *Connect Project*. *Connect* was a cluster-randomized controlled trial of the impact of community health workers on child survival that was conducted by the Ifakara Health Institute in Tanzania from 2011-2015 (57,58). *Connect* was situated in the sentinel areas of the Ifakara and Rufiji Health and Demographic Surveillance Systems (HDSS) in three rural districts (59,60). The Ifakara HDSS is in Morogoro, a landlocked region in southwestern Tanzania, and is approximately 500 km from Dar es Salaam. The Rufiji HDSS is in Rufiji district on the Indian Ocean coast approximately 150 km south of Dar es Salaam. The villages within these sites are largely agrarian and depend on subsistence agriculture, fishing, and petty trading. See Figure 1.

**Fig 1.**
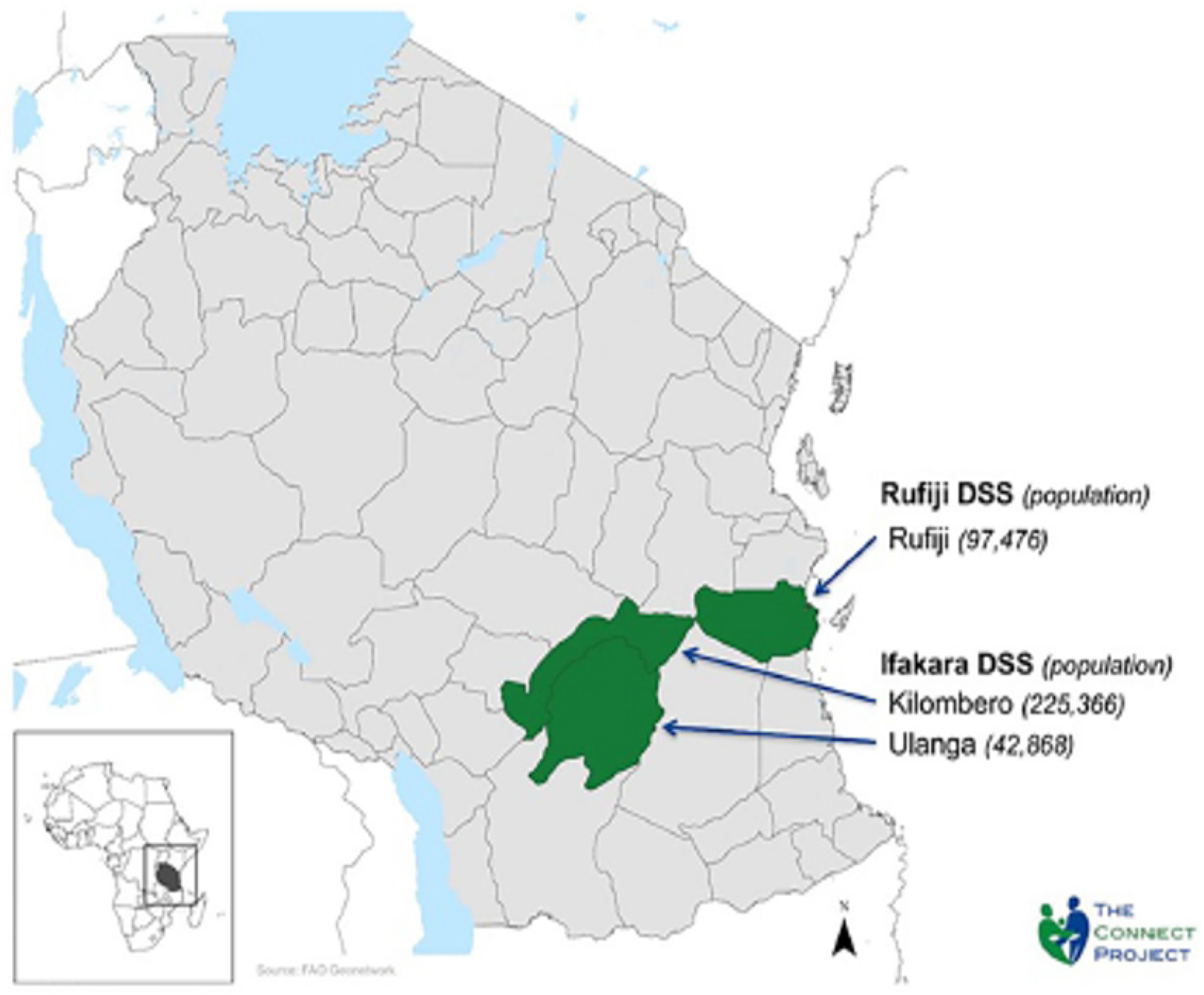
The Connect Project Study Areas.

Between April and September 2011 *Connect* conducted a household survey in 101 villages to obtain baseline measures of key MNCH behavior and service utilization indicators. At this time, *Connect* also carried out a health facility survey using the SARA tools in the 109 health facilities in the three districts to understand the context of MNCH service provision. Participants were eligible for enrollment in the household survey if they were resident in a household randomized to either study arm of the *Connect* trial, female and between the age of 18 and 49 or the caretaker of an under-five year-old child. Potential participants were identified using the census data from the HDSS and randomly selected within each village. Data collectors approached them in their homes to recruit them for the study. Health facilities were eligible for the SARA if district health authorities reported that they provided MNCH services and were in the study districts (Fig. 1). Data collectors and district health authorities visited each of these facilities to explain the study and obtain facilities’ staff cooperation with data collection. The SARA obtained information from facilities’ staff on the villages in their catchment areas. Data collectors used geographic positioning system tracking devices and mapped facility-to-village travel distances in a geographic information system database (61,62).

*Connect* leveraged the longitudinal HDSS platforms for its impact evaluation. Individuals were eligible for participation in HDSS data collection if they were residents of a household in any of the 101 villages under demographic surveillance, not a minor, and could report on the occurrence of key demographic events (births, deaths, in- and out-migrations) that took place in the household. Until December 2015, staff of both HDSS visited households in the sentinel areas every four months to collect information on household members, their relationships, ages, and sexes; births, deaths, and in- and out-migration of household members. Every 1-2 years, censuses were undertaken to enumerate old and new households. The censuses obtained data on maternal education attainment and household income and assets (63,64). For our analysis, we linked household survey, SARA and HDSS data by village name.

### Creating ‘effective coverage scales’ for specific domains of MNCH

We followed a multi-step process to develop scales of MNCH implementation strength that was exerted by PHC facilities in the *Connect* study areas on the villages that they served. For this, we used data from four modules of the SARA (excluding the fifth module which obtained data on implementation costs) and the household survey. We restricted our sample to the PHC facilities (dispensaries and health centers, not hospitals) to which residents from the 101 study villages would go for care.

The four SARA modules compiled categorical indicators on multiple domains of care: (1) general facility readiness (staffing levels, management practices and infrastructure), (2) family planning, (3) antenatal care, (4) intrapartum care, (5) postnatal care, (6) preventive services for children (e.g., immunizations, insecticide treated nets, counseling, assessment, classification components of IMCI) and (7) sick childcare (trained staff and supplies to care for respiratory illness, diarrheal disease, malaria, malnutrition). For indicators with more than one response category, we created dummy variables so that all indicators used in the analysis were binary. For all indicators in each domain, we calculated Cronbach’s alpha coefficients to establish internal consistency and found that all sets of indicators reported coefficients of 0.8 or higher (65). For each domain, we calculated ‘effective coverage indices’ using a weighted average approach. For each facility, we created the general facility readiness effective coverage index by summing the values for each of the indicators on staffing levels, management practices and infrastructure availability, respectively, and dividing each total by the number of indicators related to each of these sub-categories. We summed these averages and divided this sum by three, the number of categories in this domain.

We followed a similar procedure for the six service specific domains. Within each domain for each facility, we summed the values of indicators that fell within three common sub-categories: (i) staff training on domain-specific skills, (ii) the range and frequency with which domain specific services were available at the facility, and (iii) current availability and recent stock outs of domain-specific supplies, medicines, and equipment. We divided each of these sums by the total number of indicators related to each sub-category, then summed those averages and divided this sum by three. To incorporate coverage into this score, we identified the villages that were in each facility’s catchment area, and used data from the household survey to calculate village-level averages of met need for family planning, and, with respect to respondents’ most recent birth, receipt of ≥4 antenatal care visits, facility-based delivery, postnatal care, immunizations and, for respondents with children that had recently had diarrheal, respiratory or febrile illness, receipt of needed medications. We aggregated these averages to estimate the ‘catchment specific coverage’ for each domain. We then multiplied the six measures of ‘catchment specific coverage’ by their corresponding domain-specific scores for each facility. With this, for each facility, we obtained seven domain specific effective coverage indices.

### Combining domain specific effective coverage indices to reduce data into independent, parsimonious scales of implementation strength

To obtain a smaller set of scales of implementation strength, we used principal components analysis (PCA). We chose PCA because of its ability to reduce the highly correlated effective coverage indices into fewer orthogonal principal components (PC), or scales, that maximize the variation in the data and represent facilities’ relative position in terms of implementation strength (66). We determined the number of PC to retain in our analysis via parallel analysis. Per convention, we retained PC with an eigenvalue of greater than or equal to 1 (42). To interpret each PC, we examined the factor loadings and cosine^2^ values that were reported for each of the indicators used to formulate them. The higher loadings and cosine^2^ values indicated greater and higher-quality representation of each domain specific effective coverage index to each PC (67,68).

Next, we created scales that represented the levels of implementation strength to which each study community was exposed. For this, we grouped facilities together if SARA respondents reported that members of the same villages went to them for care or if the facility was within five kilometers of the same villages. We combined the PC values of facilities in the same group using three-level Bayesian mixed effects models with villages nested within catchment areas, and catchment areas nested within districts, and fixed effects to denote villages distance from the nearest facility and population size (69). Functionally, these models used the PC values of each facility for each retained scale as the prior distribution to produce a posterior distribution of values that represented the overall levels of implementation strength that facility groups exerted collectively (70). The benefit of this approach is that it borrows information from facilities within each group to estimate mean implementation strength scores that are shrunk to a central value, which results in more stable estimates with smaller standard errors (71).

### Estimating the association between three dimensions of implementation strength on newborn and child mortality

We merged the combined scale values that had been assigned to villages to a longitudinal dataset from the HDSS that included the survival trajectories and individual- and household level covariates for children that were born between March and November 2011. We then estimated the relationship between the implementation strength scores exerted by facility groups and the risk of child mortality, modelling the implementation strength scales in their continuous form. To address potential confounding, we incorporated fixed effects for covariates at the child level (child sex, birth order, previous and subsequent birth interval durations), mother-level (age, marital status, years of schooling), household socio-economic status (SES) ranking (1-5), and contextual variables (distance from village to nearest health facility, whether a community health worker worked in the village, HDSS zone in which villages were nested). To select our modelling strategy, we conducted the Schoenfeld test of residuals to determine whether the assumption of proportional hazards was met (72). Because multiple covariates marginally failed to satisfy this requirement, we elected to use Weibull parametric hazard regression models, which capture the underlying hazard of child mortality that is known to be high during the newborn period and decline as children age. Children’s survival durations took into account the possibility of exit from the cohort due to outmigration or death, and observations that experienced neither outcome by December 31, 2015 were subject to right censoring during analysis. To account for clustering of births within villages, we incorporated in our models a random effect for village.

In our longitudinal dataset, there was missing data for two covariates, household SES and mothers’ years of schooling, for roughly one-fourth of the sample. After examination, we determined that the omission of values reflected a ‘missingness at random’ pattern. To address this challenge, we compared three approaches: First, we omitted the two covariates from our models and conducted a complete case analysis; Second, we imputed missing values with the community-level median value for the two covariates; Third, we used multiple imputation with chain equations (MICE) (73). The results that these approaches produced were virtually identical as shown in Supplemental File 1 (S1 Appendix). Therefore, we report the results of the third approach. We conducted the entire analysis in *R Studio 4.0.2*.

Approval for this study was granted by the ethical review boards of the Ifakara Health Institute (IHI/IRB/No. 16-2010), the National Institute for Medical Research of Tanzania (NIMR/HQ/R.8a/Vol.IX/1203), and the Institutional Review Board of Columbia University Medical Center (Protocol AAF3452). To obtain the data that was used for this study, the study team first obtained information consent from human participants. Regarding the SARA and household survey, which took place from April to September 2011, to enroll subjects, data collectors trained and employed by *Connect* read an informed consent form aloud to potential participants. Those that agreed to participate signed the form or provided an inked thumbprint. A data collection team member that worked for the respective District Health Management Teams witnessed each enrollment episode and provided a countersignature. Data collection on the longitudinal cohort was managed by the HDSS teams of the Ifakara Health Institute in Kilombero and Rufiji, respectively. To ensure compliance with their mandated data collection procedures, at each follow up visit each HDSS respondent was provided with an explanation of the research purpose, interviewing process and potential uses of participants’ demographic and household information. Those that agreed to participate, then provided written informed consent under the observation of HDSS staff. This study did not enroll minors. Regarding the SARA and household survey, information that could identify individual participants with the data they provided was kept on the informed consent forms. These forms were transferred to a secure and private location at the Ifakara Health Institute as soon as possible after each episode of data collection. After forms were transferred to this location, the authors had no access to information that could identify individual participants. Datasets availed to the authors from the HDSS for this analysis were completely deidentified at the time of data transfer.

## Results

Based on information from SARA respondents and travel distances, we found that the 101 study villages sought PHC from 56 primary health care facilities. See Figure 2 and Table 1.

**Fig 2:**
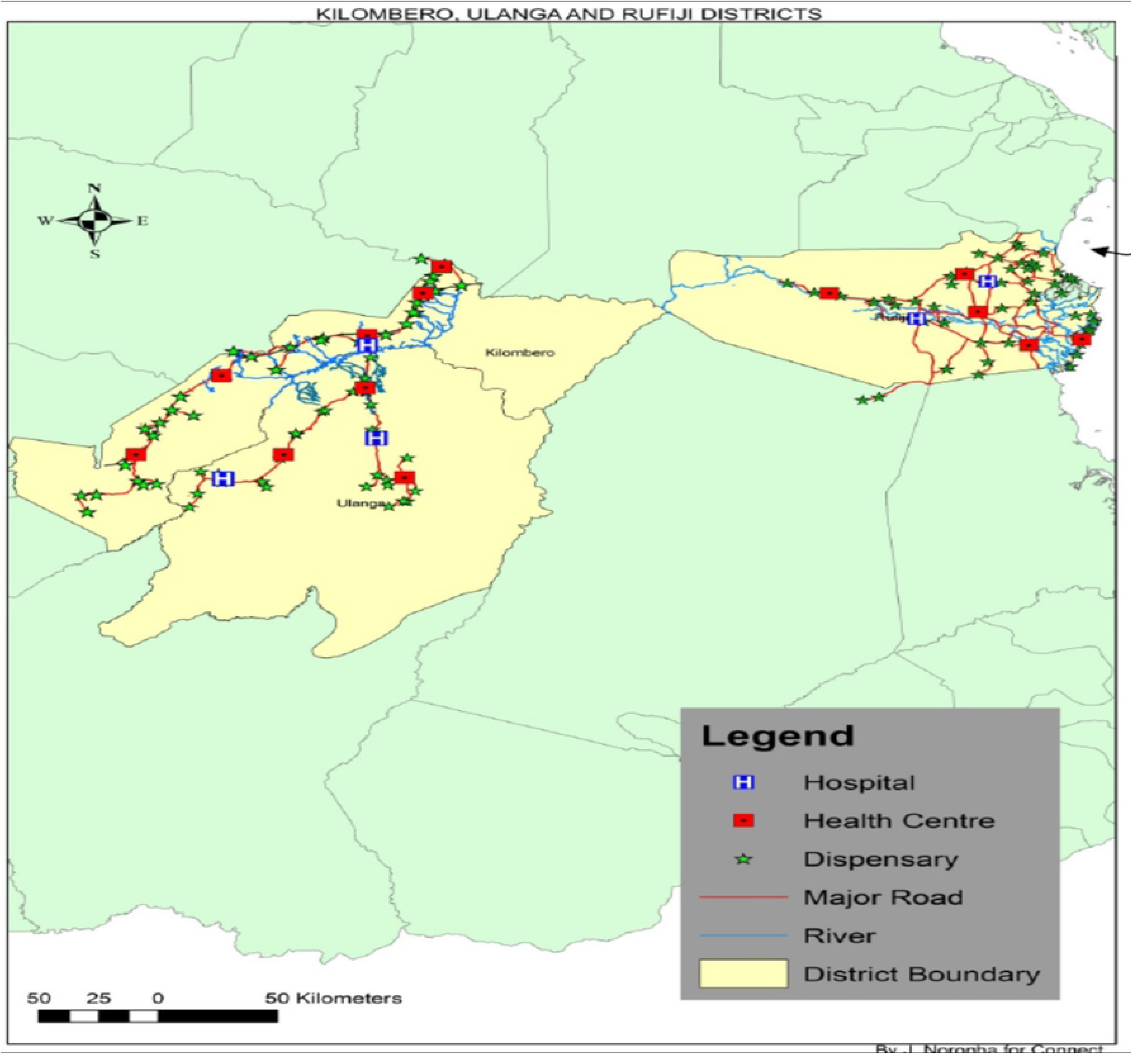
Location of health care facilities in the study area.

**Table 1:**
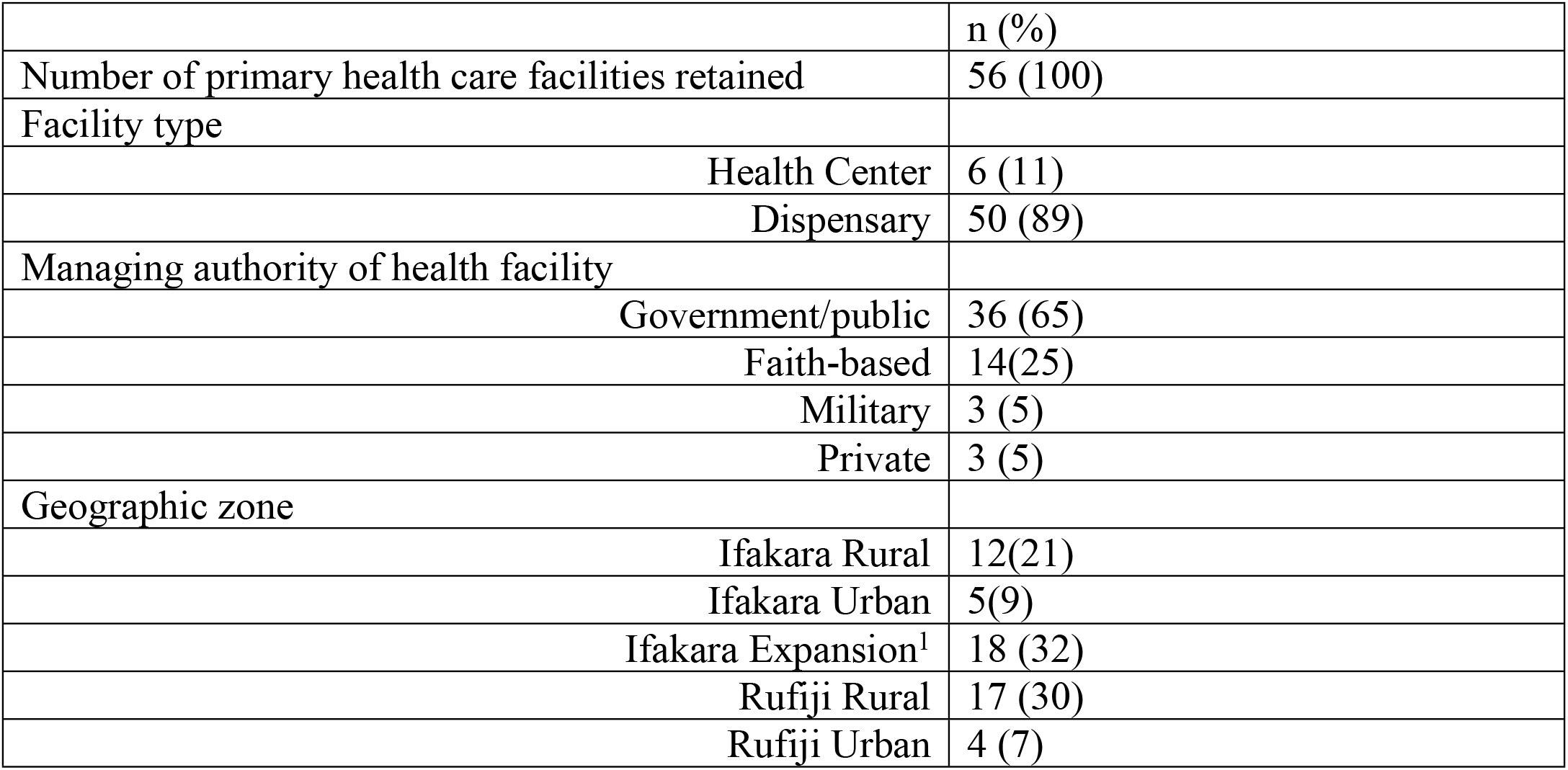
Background characteristics of primary health care facilities (n = 56)

|  | n (%) |
| --- | --- |
| Number of primary health care facilities retained | 56 (100) |
| Facility type |  |
| Health Center | 6 (11) |
| Dispensary | 50 (89) |
| Managing authority of health facility |  |
| Government/public | 36 (65) |
| Faith-based | 14(25) |
| Military | 3 (5) |
| Private | 3 (5) |
| Geographic zone |  |
| Ifakara Rural | 12(21) |
| Ifakara Urban | 5(9) |
| Ifakara Expansion<sup>1</sup> | 18 (32) |
| Rufiji Rural | 17 (30) |
| Rufiji Urban | 4 (7) |

In total, 234 indicators from the SARA were used to create domain specific effective coverage indices (47 indicators on general facility readiness, 23 family planning services, 22 antenatal care, 54 delivery services, 23 postnatal care, 31 on preventive childhood services, 34 on sick child care). Table 1 presents descriptive data on the domain specific effective coverage indices calculated for the facilities in our sample. Of the 56 facilities, 6 were health centers, and 50 were dispensaries.

Thirty-five of the facilities were in the sentinel areas of the Ifakara HDSS (12 in Ifakara Rural, 5 Ifakara Urban and 18 Ifakara Expansion) and 21 in the Rufiji HDSS (17 in Rufiji Rural, 4 Rufiji Rural). Supplemental File 2 contains a table that presents the median and range of scores of the effective coverage indices by HDSS zone (S2 Appendix).

Supplemental File 3 shows the Scree plot that was produced by the PCA and guided our parallel analysis (S3 Appendix). The first three PC reported eigenvalues of 1 or greater and together accounted for 77% of the variance among the seven domain specific effective coverage indices, whereas the remaining four scales held appreciably less explanatory potential. Thus, we chose to retain the first three scales only.

Figures 3a and 3b reflect the factor loadings and cosine^2^ values for each PC, which reflect the relative contribution of each underlying effective coverage index to the retained scales. We found that the first PC, which accounted for 46% of variation in the effective coverage indices, was an implementation strength scale that reflected facilities’ relative position in terms of the availability and readiness to provide preventive health services, mostly child-related, with high loadings and cosine^2^ values for general facility readiness, antenatal care, postnatal care, and preventive childhood services. The second PC, which accounted for 18% of the variation, represented a scale of facilities’ readiness to provide sick childcare and, to a lesser extent, family planning services; and the third PC, which accounted for 13% of the variation, represented a scale of facilities’ readiness to provide intrapartum care.

**Fig 3a and 3b:**
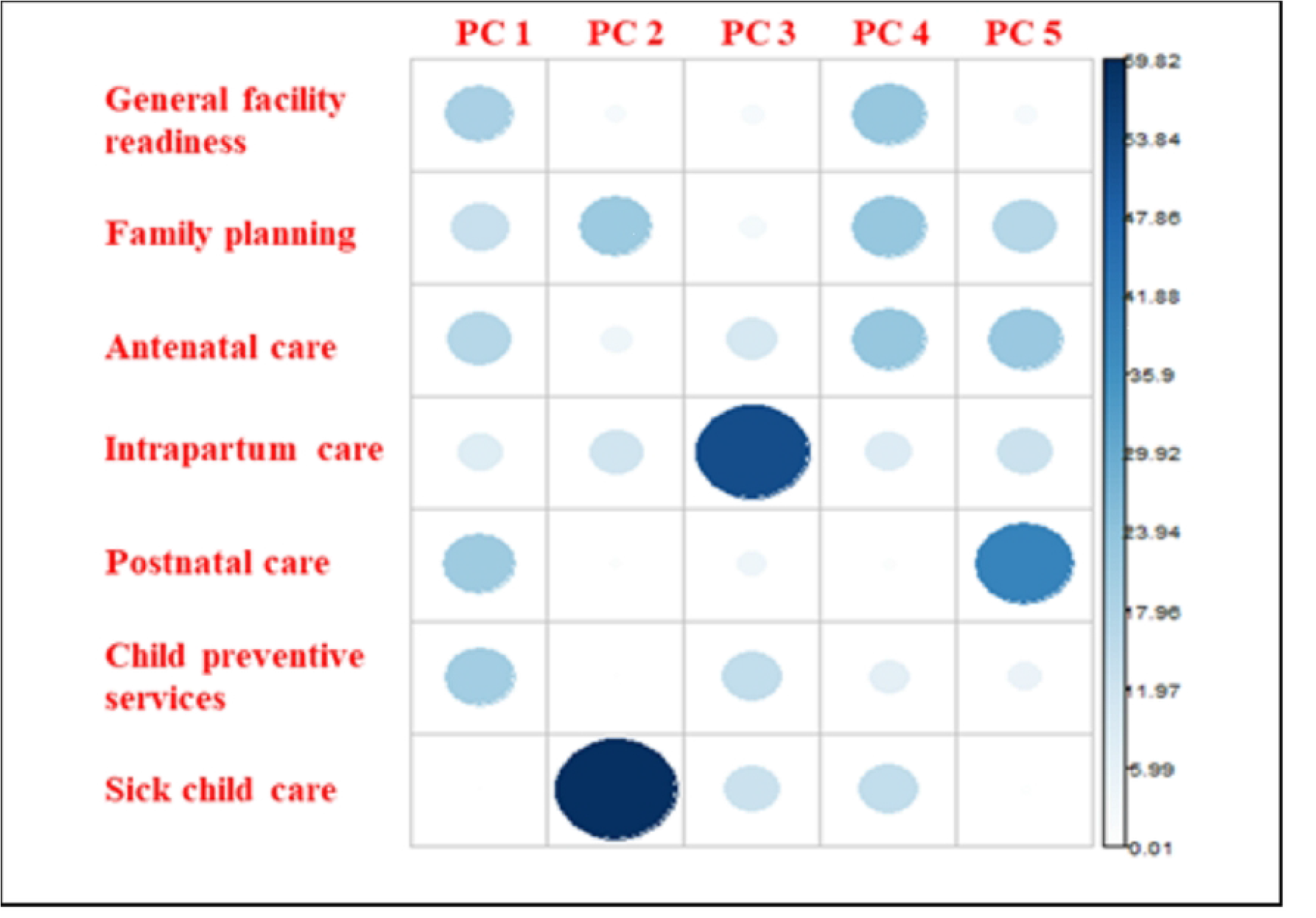
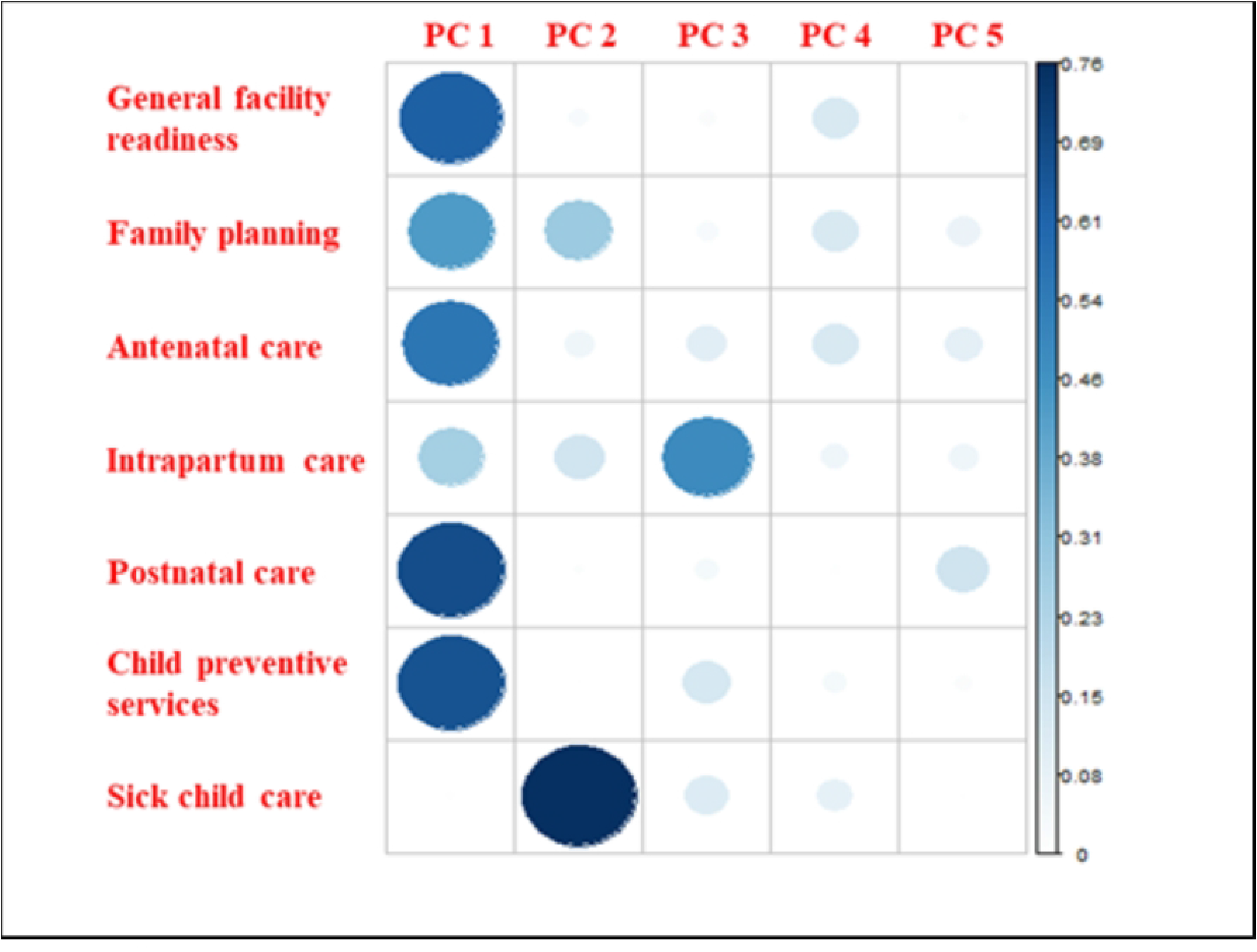
Correlation plot of factor loadings (3a) and Cosine^2^ values (3b) produced by PCA of MNCH effective coverage indices.

We used information provided by SARA respondents and geographic data to determine which of the 56 facilities in our sample served the 101 villages in the study area respectively. In doing so, we identified 89 different combinations of facilities. On average, villages were within the catchment areas of three primary care facilities (range: 1, 7). For each village, we computed the collective implementation strength that was exerted on them by the group of facilities in whose catchments they were located using a Bayesian mixed effects model and estimating the empirical Bayes mean implementation strength scores for the three scales that we retained. Figures 4a-c illustrates the distribution of implementation strength scores assigned to the 89 different facility groups that defined the catchment areas in which the 101 study communities were nested.

**Fig 4a-c:**
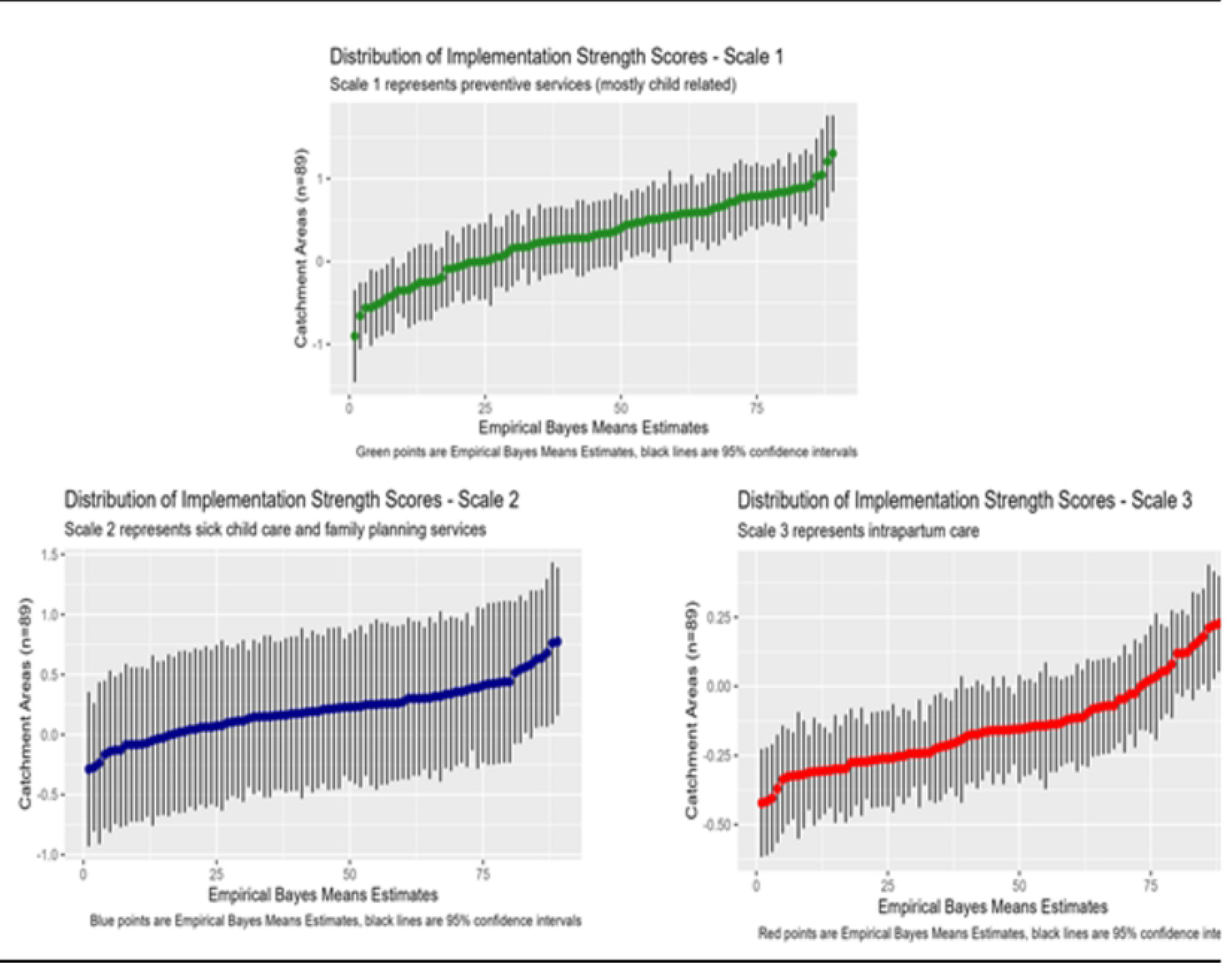
Distribution of implementation scores across primary health care facility catchment areas in the study area (n = 89)

Supplemental Files 4a-c illustrates the geographic variation in implementation strength, comparing the distribution of implementation strength scores between groups of villages that were in five administrative zones within the study area, three in the Ifakara HDSS (Ifakara rural, urban and expansion areas) and two in Rufiji (rural and urban) (S4 Appendix). We observe the greatest variation in median implementation strength scores among zones for the first scale, which represents the availability and readiness to provide preventive care services for newborns and children as well as antenatal care, and the least variation between zones for the second scale, which represents access to sick childcare and family planning services.

Table 2 describes the characteristics of 8,999 children that were born from March-November 2011, the period spanning approximately three months before and after the start and end of the SARA, in the villages whose residents report to the above-described 56 facilities for care. Among these children, 526 (5.8%) died before December 31, 2015, when the follow up of the cohort ended. The median period of observation of children in the cohort was 46 months (range: 0, 58).

**Table 2:**
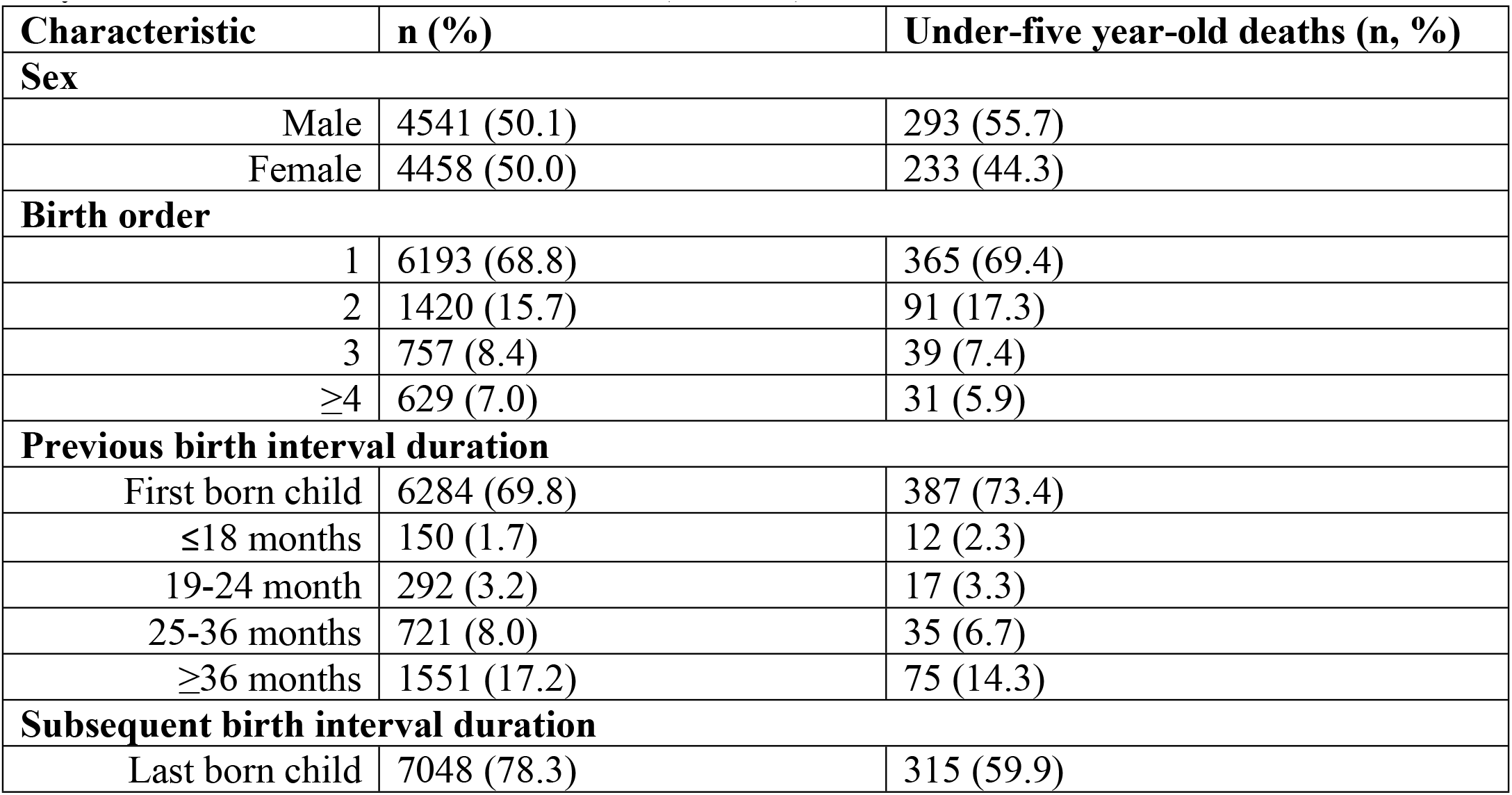
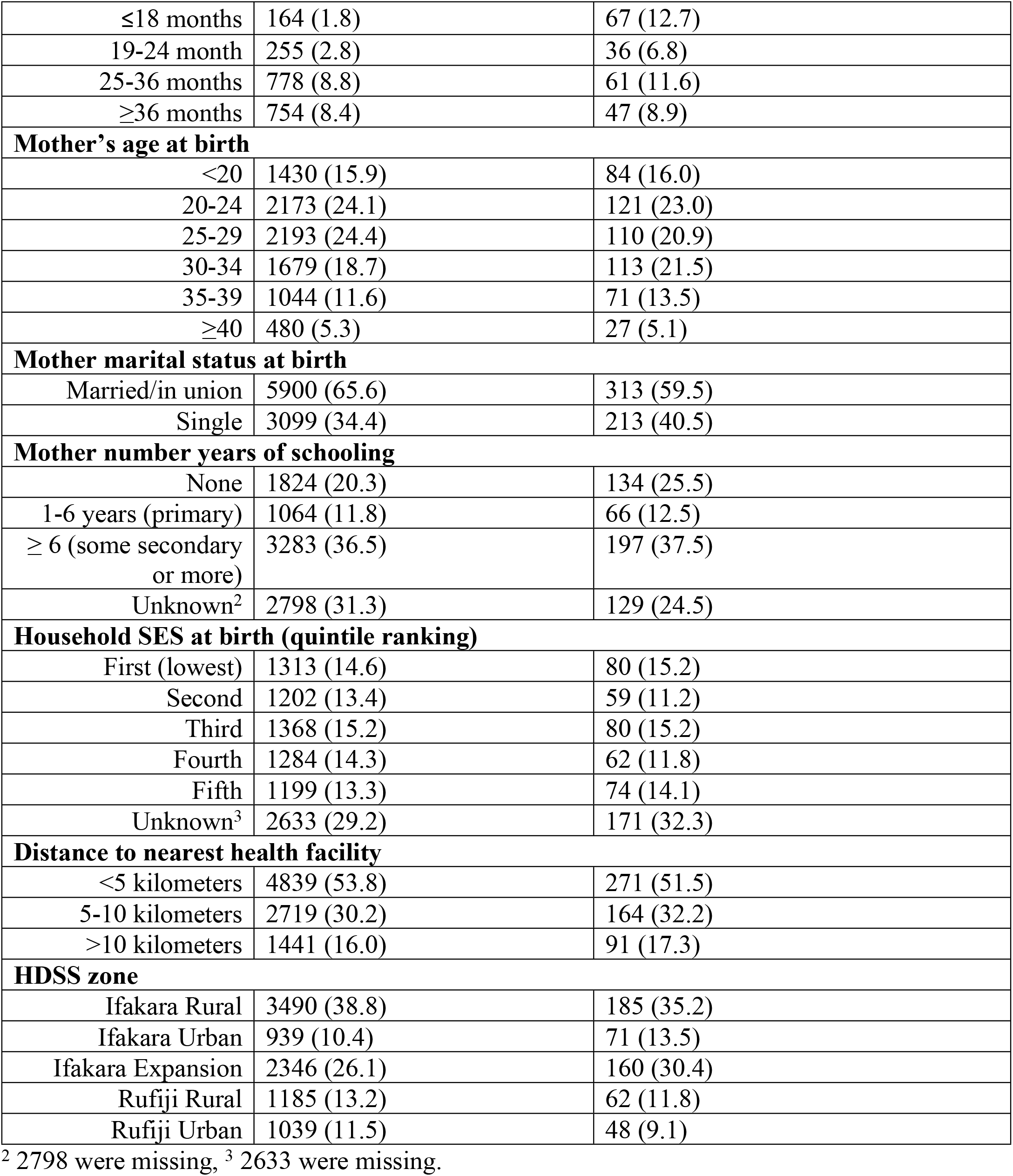
Characteristics of children born in catchment areas of the 56 primary health care facilities in the study area between March and November 2011 (n = 8,999)

| Characteristic | n (%) | Under-five year-old deaths (n, %) |
| --- | --- | --- |
| **Sex** |  |  |
| Male | 4541 (50.1) | 293 (55.7) |
| Female | 4458 (50.0) | 233 (44.3) |
| **Birth order** |  |  |
| 1 | 6193 (68.8) | 365 (69.4) |
| 2 | 1420 (15.7) | 91 (17.3) |
| 3 | 757 (8.4) | 39 (7.4) |
| ≥4 | 629 (7.0) | 31 (5.9) |
| **Previous birth interval duration** |  |  |
| First born child | 6284 (69.8) | 387 (73.4) |
| ≤18 months | 150 (1.7) | 12 (2.3) |
| 19-24 month | 292 (3.2) | 17 (3.3) |
| 25-36 months | 721 (8.0) | 35 (6.7) |
| ≥36 months | 1551 (17.2) | 75 (14.3) |
| **Subsequent birth interval duration** |  |  |
| Last born child | 7048 (78.3) | 315 (59.9) |
| ≤18 months | 164 (1.8) | 67 (12.7) |
| 19-24 month | 255 (2.8) | 36 (6.8) |
| 25-36 months | 778 (8.8) | 61 (11.6) |
| ≥36 months | 754 (8.4) | 47 (8.9) |
| **Mother's age at birth** |  |  |
| <20 | 1430 (15.9) | 84 (16.0) |
| 20-24 | 2173 (24.1) | 121 (23.0) |
| 25-29 | 2193 (24.4) | 110 (20.9) |
| 30-34 | 1679 (18.7) | 113 (21.5) |
| 35-39 | 1044 (11.6) | 71 (13.5) |
| ≥40 | 480 (5.3) | 27 (5.1) |
| **Mother marital status at birth** |  |  |
| Married/in union | 5900 (65.6) | 313 (59.5) |
| Single | 3099 (34.4) | 213 (40.5) |
| **Mother number years of schooling** |  |  |
| None | 1824 (20.3) | 134 (25.5) |
| 1-6 years (primary) | 1064 (11.8) | 66 (12.5) |
| ≥ 6 (some secondary or more) | 3283 (36.5) | 197 (37.5) |
| Unknown<sup>2</sup> | 2798 (31.3) | 129 (24.5) |
| **Household SES at birth (quintile ranking)** |  |  |
| First (lowest) | 1313 (14.6) | 80 (15.2) |
| Second | 1202 (13.4) | 59 (11.2) |
| Third | 1368 (15.2) | 80 (15.2) |
| Fourth | 1284 (14.3) | 62 (11.8) |
| Fifth | 1199 (13.3) | 74 (14.1) |
| Unknown<sup>3</sup> | 2633 (29.2) | 171 (32.3) |
| **Distance to nearest health facility** |  |  |
| <5 kilometers | 4839 (53.8) | 271 (51.5) |
| 5-10 kilometers | 2719 (30.2) | 164 (32.2) |
| >10 kilometers | 1441 (16.0) | 91 (17.3) |
| **HDSS zone** |  |  |
| Ifakara Rural | 3490 (38.8) | 185 (35.2) |
| Ifakara Urban | 939 (10.4) | 71 (13.5) |
| Ifakara Expansion | 2346 (26.1) | 160 (30.4) |
| Rufiji Rural | 1185 (13.2) | 62 (11.8) |
| Rufiji Urban | 1039 (11.5) | 48 (9.1) |

<sup>2</sup> 2798 were missing, <sup>3</sup> 2633 were missing.

Table 3 presents the results of our analysis of the relationship between the three scales which represent the implementation strength of MNCH to which children in the study area were exposed during early infancy in 2011 and the risk of their dying before December 31, 2015. The first model is an unadjusted analysis of the effect of the three dimensions of implementation strength on child mortality, the second the same analysis however adjusting for individual mother- and child-level covariates, and the third the same analysis however adjusting for individual mother- and child-level and contextual covariates. From our analysis, we observe an association between the first scale, which gauges the strength of general facility readiness and antenatal, postnatal, and preventive early childhood health care services accessible to the population, and the risk of dying during childhood. After adjustment for potentially confounding factors, the third model reports that for each unit increase in the implementation strength of these interventions, the risk of dying during childhood is 0.62 times lower (HR: 0.62; 95% CI: 0.40, 0.97).

**Table 3:** Associations between child mortality risk and implementation strength scores reported from Model 1 (adjusted, scores 1-3 only), Model 2 (scores and mother- and child-level covariates), and Model 3 (scores and mother- and child-level covariates and contextual-level covariates) (n = 8,999)

|  | **Model 1** |  | **Model 2** |  | **Model 3** |  |
| --- | --- | --- | --- | --- | --- | --- |
|  | HR | 95% CI | HR | 95% CI | HR | 95% CI |
| **Implementation strength scores** |  |  |  |  |  |  |
| Score 1 (ANC, PNC, and preventive child care) | 0.76 | 0.60, 1.00 | 0.70* | 0.54, 0.90 | 0.62* | 0.40, 0.97 |
| Score 2 (sick child care) | 0.74 | 0.43, 1.25 | 0.60* | 0.35, 0.99 | 0.56* | 0.31, 0.99 |
| Score 3 (intrapartum care) | 0.86 | 0.43, 1.73 | 0.62 | 0.31, 1.23 | 0.47 | 0.19, 1.22 |
| **Child sex** |  |  |  |  |  |  |
| Female | - | - | - | - | - | - |
| Male | - | - | 1.25* | 1.05, 1.48 | 1.25* | 1.05, 1.49 |
| **Birth order** |  |  |  |  |  |  |
| No. children (cont.) | - | - | 0.99 | 0.88, 1.13 | 1.00 | 0.88, 1.13 |
| **Previous birth interval** |  |  |  |  |  |  |
| Months (cont.) | - | - | 1.00 | 0.99, 1.00 | 1.00 | 0.99, 1.01 |
| **Subsequent birth interval** |  |  |  |  |  |  |
| Months (cont.) | - | - | 1.01*** | 1.00, 1.02 | 1.01*** | 1.00, 1.02 |
| **Mother age at birth** |  |  |  |  |  |  |
| Years (cont.) | - | - | 1.01 | 0.99, 1.02 | 1.01 | 1.00, 1.02 |
| **Mother marital status at birth** |  |  |  |  |  |  |
| Married/in union | - | - | - | - |  |  |
| Single | - | - | 1.44** | 1.20, 1.72 | 1.43** | 1.19, 1.72 |
| **Mother number years of schooling** |  |  |  |  |  |  |
| Year of schooling (cont.) | - | - | 0.97<sup>□</sup> | 0.94, 1.00 | 0.97 | 0.95, 1.01 |
| **Household SES at birth (quintile ranking)** |  |  |  |  |  |  |
| Fifth | - | - | - | - | - | - |
| Fourth | - | - | 0.78<sup>□</sup> | 0.60, 1.02 | 0.79<sup>□</sup> | 0.60, 1.03 |
| Third | - | - | 0.96 | 0.74, 1.26 | 0.99 | 0.75, 1.29 |
| Second | - | - | 0.92 | 0.70, 1.22 | 0.95 | 0.71, 1.26 |
| First | - | - | 0.91 | 0.68, 1.21 | 0.93 | 0.70, 1.25 |
| **Distance to nearest health facility** |  |  |  |  |  |  |
| Kilometers (cont.) | - | - | - | - | 1.14<sup>□</sup> | 0.98, 1.32 |
| **HDSS zone** |  |  |  |  |  |  |
| Ifakara Expansion | - | - | - | - | - | - |
| Ifakara Rural | - | - | - | - | 0.85 | 0.58, 1.22 |
| Ifakara Urban | - | - | - | - | 1.25 | 0.81, 1.95 |
| Rufiji Rural | - | - | - | - | 1.12 | 0.70, 1.81 |
| Rufiji Urban | - | - | - | - | 0.95 | 0.54, 1.68 |
| **Community Health Worker Deployed to Community** |  |  |  |  |  |  |
| Yes |  |  |  |  | 0.93 | 0.76, 1.13 |

□ = p-values < 0.1, \* = p-values < 0.05, \*\* = p-values < 0.01, \*\*\* = p-values < 0.001

The third model suggests a more protective effect of implementation strength on child mortality with regards to the second scale, which represents the availability and readiness of nearby facilities to provide sick child care. After adjustment for multi-level covariates, we noted that among children in our cohort, increases exposure to higher levels of sick child care implementation intensity were associated with, on average, with 0.56 times lower risk of dying during childhood (HR: 0.56, 95% CI: 0.31, 0.99). There was no statistically significant association detected between variation in the third scale of implementation strength, which represents the strength of intrapartum care services available to the population, and child mortality risk. In addition to evaluating the child mortality response, we adapted Models 1-3 so that they could indicate whether variation in implementation strength, as represented by the three scales, were associated with children’s risk of dying in the first month and first year of life, respectively. These analyses reported hazards ratios of magnitudes that were similar to those reported by Model 3, but these results were not statistically significant (S5 Appendix).

## Discussion

Our analysis reports that children exposed to higher levels of implementation strength of preventive and curative childcare, including antenatal care, postnatal care, and family planning services available to mothers, from the PHC facilities that served their community at the time of birth and early infancy were less likely to die during childhood than those exposed to lower levels of implementation strength. Whereas the same analyses of the relative risk of newborn and infant mortality associated with implementation strength did not reveal a significant relationship, we believe that this owes the limitations in the power accorded by the sample size and number of mortality events in the first month and year of life in our cohort. Furthermore, we observe no association between child mortality and the third implementation strength scale, which represents the strength of intrapartum care services. This finding contradicts similar studies which measured the health response to obstetric care quality. For example, Tiruneh et al. (2018). in Ethiopia used an additive approach to develop a gradient of EmONC implementation strength and conducted an analysis which reported a positive association between higher levels of EmONC implementation strength and facility-based delivery and met need for EmONC (34). Although our finding may be surprising, it is important to recall that the third scale explained a relatively small proportion of the overall variance among the effective coverage indices that we reduced into our independent gradients of implementation strength. If, in our analysis of SARA data, we had included hospitals and higher-level health centers, which are relatively better equipped to handle labor and delivery, it is likely that the sample would have contained more variation with respect to this domain of care. In turn, this might have led to a set of scales that demonstrated a more potent effect of intrapartum care implementation strength.

Our findings contribute to the debate about the role of primary health care programs and services in the precipitous child mortality decline that Tanzania experienced from 2000-2015. Importantly, they corroborate the findings issued by Masanja et al. (2008) in their analysis of the trends and drivers of child mortality using four consecutive DHS surveys between 1990 and 2008. This study identified the synergistic effect of increases in public sector health expenditure, implementation of decentralization policies and expansion in the coverage of high impact child health interventions, such as insecticide treated nets (ITN), extended program immunizations (EPI), and IMCI, as contextual factors important to the reduction of child mortality observed in Tanzania during this period (74). Afnan-Holmes and coauthors, make similar observations when reviewing Tanzania’s achievement of Millennium Development Goal 4, also attributing success to increased coverage of EPI, IMCI and ITN (75). Our analysis underscores the importance of investments in expanding these interventions. Furthermore, our interpretation of the first set of factor loadings produced by the PCA suggests that ensuring effective coverage of routine antenatal and postnatal care as well as general facility readiness, in terms of regular staffing, general infrastructure and routine management inputs and processes, may be as important to enhancing child survival prospects in the population as preventive and promotive health interventions that target children under five. This information is relevant to health system managers and planners in Tanzania as they prioritize ways to invest limited resources in ensuring effective coverage of PHC services and scaling up.

This analysis illuminated an appreciable child mortality response to slight variation in the strength of PHC performance within small geographic areas. In our examination we distinguished the relative contributions of different domains of MNCH to this variability and provided insight on how improvements vis-à-vis these domains can impact children’s prospects of surviving childhood. Yet, we did not explore the factors that explain why, within relatively small areas, some facilities perform better than others. Previous studies in Tanzania have sought to understand how differences between nearby facilities condition patient care seeking. For example, one study reported that care seeking behaviors were shaped by differences between facilities in terms of the quality of consultations and availability prescriptions, knowledge-level of staff, and availability of physicians and essential supplies and equipment (76). Another study sought to understand the drivers of variation between nearby facilities in rural areas in terms of provider competence and practice quality, and reported that this was a function of facility ownership (private for profit, non-governmental organization, public sector), population density of facilities’ catchment area, and health workers training, tenure and experience (77). Other studies in Tanzania on this topic have focused on specific domains of MNCH. Kahabuka et al. (2011) attributed patient care seeking for preventive and sick child care services to differences between nearby facilities in terms of the availability of diagnostic equipment, essential medicines and skilled staff (78). Kanté et al. (2016) reported that differences between facilities’ performance of emergency obstetric care signal functions conditioned women’s care seeking for intrapartum care (79). Future research, in Tanzania and similar settings, should continue to examine the determinants and processes that generate within-small area variation in service readiness, availability and quality since this information, as our findings suggest, could help address problems that underlie preventable child deaths.

Effective coverage measures that link population level data on access to health care and health outcomes with facility data on health care quality are increasingly reported (80–82). However, there is little guidance on appropriate methods for linking, e.g., which data to link, for which units and with what temporal alignment (46). Furthermore, decisions to link data sets are usually not foreseen during study design, and, therefore, researchers must be opportunistic in the linkage methods they employ. In our analysis, we linked individual-level data on child survival to summary scores of MNCH coverage effectiveness of the facilities in the environs of children’s household and communities and found that the contextual effect of implementation strength in these areas was associated in lower risks of child mortality. However, this might differ from an analysis of the same relationship that linked individual outcomes with performance scores of exact facilities where children obtained care. Future research is needed on effective coverage estimation and its health effects that compares methods that operationalize coverage in geographic terms versus in terms of where individuals utilized services. Identification of biases associated with either linkage approach will help health systems researchers determine the data and data systems requirements for measuring health systems strengthening and its impacts.

To our knowledge this is the only study that has benefitted from the ability to link data on ‘dose delivered’ of implementation strength within local health systems with longitudinal data on the survival of children nested in the underlying population. Although other studies, for example in Ethiopia and Malawi, addressed similar questions related to implementation strength of MNCH services, the larger geographic scope of their analysis and lack of prospective data on the outcome compelled authors to draw upon repeat cross-sectional data from the DHS or special project surveys, and perform ecological analyses, which are subject to biases (38,44). Whereas these findings are valuable insofar as they evaluate the impact of the large-scale rollout of national child survival programs, our analysis fills an important gap in that it reports the effects of variation in dose delivered of implementation strength by routine delivery systems within a relatively small area on individual level mortality risks that unfold over time after exposure. Future research should seek opportunities to leverage existing longitudinal data collection platforms and integrate them into investigations of the population-level health response to changes in health systems strength.

Our analysis is not without limitations. First, though we opted to use PCA as a data reduction approach, we recognize that this approach has drawbacks. For example, there are examples that have demonstrated that the use of PCA results in the misclassification of subjects vis-à-vis the gradient of the underlying construct when the PCs used explained less than 30% of total variance (83). Although our finding that higher levels of readiness and availability of sick child care was associated with lower child mortality risk seems intuitive, it is not immune to this type of critique. Second, despite our review of the risks of weighted-additive methods, we used that approach to derive effective coverage indices of different domains of MNCH that we later subjected to PCA. This came after reviewing alternative data reduction strategies, including use of PCA only to reduce the 234 indicators in the SARA to implementation strength scales. In the end, we felt that the weighted-additive approach, enabled us to represent the natural partition in our data between domain-specific measures of MNCH availability and readiness and best identify which components of MNCH care are most relevant to child mortality reduction. Third, there were significant levels of missingness for two covariates in the HDSS, which supplied our data on child survival. However, after comparing three approaches for addressing this problem (complete case analysis, median imputation, and MICE) we found that comparable results were obtained under all strategies, which indicated that missing data did not seriously affect the overall association between implementation strength and child survival. Fourth, the analysis assumes that the levels of implementation strength that we obtained from the cross-sectional SARA from May to September 2011 reflect the longer period for which we subset the longitudinal data from the HDSS (child survival trajectories starting at birth from March to November 2011). Fifth, our analysis links children born during the period surrounding the SARA with data on the facilities in whose catchment their communities were located; however, our analysis could not ascertain whether these children ever sought health care at other health facilities during their first 4-5 years of life. Finally, although the analysis was able to link within district variation in levels of implementation strength exerted by primary health care facilities with longitudinal data on child survival, these data are observational and, thus, fall short of permitting inference that is truly causal.

## Conclusion

We developed gradients that quantify the strength with which MNCH services were delivered to the populations in the local health systems in three districts of Tanzania in 2011, and, by linking these scales with longitudinal survival trajectories of children born in the underlying population, we evaluated whether variation in implementation strength at the time of childbirth and early infancy were associated with mortality risk during childhood. The results suggest that the intensity with which preventive care services, including general facility readiness, antenatal and postnatal care, and preventive care for children, were made available to the population, as well as that of curative care and family planning services, was associated with lower mortality risk. Since these scales reflect the performance of interventions that are part of the Tanzania’s essential MNCH services package through routine delivery system channels, local health authorities can use these metrics, which were derived by using data readily available from facilities, to better understand the health impact of their implementation on the populations they serve. Additional research of this nature should be undertaken using data from hospitals to obtain data with greater variation on the performance of obstetric and gynecological services to better understand how health systems meet the needs of women during the crucial intra- and postpartum periods. Health system managers and decision-makers can use this information to inform planning, resource allocation and implementation adjustments and maximize the impact of health system strengthening on population health.

## Data Availability

Data cannot be shared publicly but can be made available subject to the approval of a material transfer agreement between parties interested in obtaining the data and the National Medical Research Coordinating Committee of the National Institutes of Medical Research of Tanzania. The MTA and application instructions are available at: https://healthresearchwebafrica.org.za/files/MTA.pdf). Those interested in obtaining the data should also contact the corresponding author for assistance with the Material Transfer Agreement.

## Supplemental information

1. Supplemental File 1 (S1 Appendix): Association between child mortality risk and MNCH implementation strength using three approaches for addressing missing data for covariates on household SES and mothers’ years of schooling. Model 1: Complete case analysis omitting covariates with missing data (n=6,068). Model 2: Addresses missing data by imputing community-level median values of household SES and mothers’ years of schooling where data are missing (n=8,999). Model 3: Addresses missing data by using multiple imputation with chained equations (n=8,999).
2. Supplementary File 2 (S2 Appendix): Effective coverage indices (median and range) for primary health care facilities (n = 56).
3. Supplemental File 3 (S3 Appendix): Scree plot reported by the principal components analysis of effective coverage indices.
4. Supplemental Files 4a (S4a Appendix): Comparison of the distribution of implementation strength scale 1’s values across geographic zones in the study area.
5. Supplemental Files 4b (S4b Appendix): Comparison of the distribution of implementation strength scale 2’s values across geographic zones in the study area.
6. Supplemental Files 4c (S4c Appendix): Comparison of the distribution of implementation strength scale 3’s values across geographic zones in the study area.
7. Supplemental File 5 (S5 Appendix): Associations between newborn (<1 month), infant (<12 months) and child mortality (<60 months) risks and implementation strength scores reported from Model 3 (IS scores, mother-, child- and contextual-level covariates) (n=8,999)

## Acknowledgments

We gratefully acknowledge the funding and advisory support of the Doris Duke Charitable Foundation (grant number DDCF2009058a) and Comic Relief UK (grant number 112259), and the contributions of staff of the Health and Demographic Surveillance Systems of the Ifakara Health Institute including Amri Shamte, Matthew Alexander, Francis Levira, Eveline Geubbles, Rose Nathan. We also recognize the contributions of Jitihada Baraka, Ruth Wilson, Awena Malendo, Mustafa Ngozi and Ahmed Hingora. Finally, we recognize with gratitude the leadership and importance of staff and supporters from Kilombero, Rufiji and Ulanga districts, including staff of their respective Council Health Management Teams and authorities of the villages in which this research was conducted.

## Footnotes

1 Expansion refers to the geographic area comprising of communities in which the Ifakara HDSS was expanded in 2010.

